# High Prevalence of *SOD1* Pathogenic Variants in the UK Biobank: Implications for Early Intervention in ALS

**DOI:** 10.1101/2025.06.12.25329421

**Authors:** Delia Gagliardi, Chiara Villella, Matteo Zanovello, Virginia Iacobelli, Stefania Corti, Giacomo Pietro Comi, Pietro Fratta, Henry Houlden, Arianna Tucci, Dario Ronchi

## Abstract

*SOD1* is the second most mutated gene in European amyotrophic lateral sclerosis (ALS) patients, after *C9orf72* expansion. Given the recent authorisation of SOD1-directed antisense oligonucleotide for use in SOD1-ALS patients, prompt screening for *SOD1* mutations in ALS patients is highly recommended. Large-scale genomic analysis could inform on the population-based prevalence of *SOD1* mutation carriers, who would potentially benefit from treatment. We aim to determine the number of people with pathogenic *SOD1* variants in the UK Biobank (UKB), to address a critical gap between clinical and genetic prevalence of SOD1-ALS, with important implications for early therapeutic intervention in presymptomatic carriers.

We analysed variants in the SOD1 gene within exome sequencing data from 470,000 individuals over 40 years old at the time of recruitment. We evaluated their pathogenic role using referenced databases and according to ACMG guidelines. Leveraging the carrier frequency in UKB and age at onset distribution data, we estimated the disease prevalence of *SOD1*-ALS.

We identified 122 individuals with monoallelic *SOD1* coding variants. 93.4% of them were asymptomatic at enrollment. In addition, the low penetrance disease allele p.Asp91Ala was observed in heterozygosis in 535 subjects, while it was never found in homozygosis. Based on this data, the expected number of people developing SOD1-ALS in the UK population, excluding the p.Asp91Ala allele, is 1.1:100,000, four times higher than the prevalence derived from clinical data.

Incomplete and age-related penetrance and heterogeneous phenotypic expression likely account for the reduced number of symptomatic patients identified. Our findings highlight the need for systematic genetic screening in the general population, which could dramatically expand the pool of individuals who might benefit from novel molecular-silencing therapies presymptomatically. Given the existence of a therapeutic option, an in-depth follow-up of these subjects is highly recommended.

## Introduction

In 1993, pathogenic variants in the superoxide dismutase 1 (*SOD1*) gene were identified as the first genetic cause of ALS, marking a pivotal moment in understanding the disease’s molecular basis^1^. This discovery initiated a 30-year journey from gene identification to the development of targeted gene therapies, creating a pathway toward potential disease prevention and highlighting the critical importance of identifying at-risk individuals before symptom onset.

*SOD1*-ALS accounts for approximately 1-6% of ALS cases globally, with significant geographic variation in prevalence, representing approximately 1-2% of sporadic cases (sALS) and up to 20% of familial forms (fALS)^2^. Clinical presentations of *SOD1*-ALS are remarkably heterogeneous, with a median age of onset around 47-48 years, variable disease progression rates, and predominantly spinal rather than bulbar onset of symptoms^3^. Penetrance of *SOD1*-ALS is incomplete and varies across different mutations^4^, ranging from almost full penetrance in the aggressive p.Ala5Val variant to a polymorphic frequency in some populations for the heterozygous p.Asp91Ala. A recent study estimated a penetrance for *SOD1*-ALS of 54% at a population level^5^.

Over 234 different variants, unevenly distributed across the five exons of *SOD1,* have been reported^6^. Despite efforts in assessing the pathogenicity of *SOD1* variants^7^, there are no consensus criteria to definitely prove their detrimental role. The vast majority of them are missense variants; however, more than 30 nonsense pathogenic variants have been reported^4^. Pathogenic variants are usually heterozygous; however, for some variants (p.Ser69Pro, p.Leu85Phe, p.N87Ser, p.Asp91Ala, p.Leu118Val, p.Val120Leu, p.Leu127Ser, p.Leu145Ser, and p.Gly28delGGACCA), a recessive inheritance has been reported^8–10^.

Biallelic variants in p.Asp91Ala, along with compound heterozygosity between the D91A monoallelic variant and other *SOD1* variants, are associated with fALS in Scandinavia. Despite being described in a heterozygous state in some ALS patients^3^, the pathogenic role of the p.Asp91Ala variant is still debated. The absence of co-segregation with the heterozygous p.Asp91Ala carrier in ALS pedigrees, the normal enzymatic activity and the presence of only minimal SOD1 microaggregates in post-mortem brains, similarly to those observed in other neurodegenerative disorders, question the pathogenicity of the heterozygous p.Asp91Ala variant^11^.

Recent therapeutic advances have targeted the toxic gain-of-function mechanism through gene silencing approaches. Tofersen, an antisense oligonucleotide that reduces SOD1 protein expression by inducing RNase H-mediated degradation of SOD1 messenger RNA, received accelerated approval from the FDA based on its ability to reduce plasma neurofilament light chain (NfL) concentrations, a biomarker reasonably likely to predict clinical benefit^12^. Moreover, longitudinal follow-up of pre-symptomatic *SOD1* variant carriers has revealed that plasma NfL concentrations rise 6-12 months before phenoconversion to clinically manifest ALS^13^. This finding has enabled the design of preventive trials (ATLAS, NCT04856982), which aim to prevent or delay the onset of clinical symptoms by administering tofersen when NfL levels rise above a predefined threshold in pre-symptomatic carriers of highly penetrant *SOD1* variants.

*SOD1*-ALS is currently the only genetic ALS form for which a molecular therapy is available, and a clinical study on presymptomatic individuals carrying aggressive variants is ongoing^13^. Moreover, *SOD1*-ALS clinical manifestation can be subtle and slowly progressing, resulting in a substantial delay (months or even years) in diagnosis^14^. Thus, identifying potential carriers in the general population represents an unprecedented opportunity to intervene before irreversible neurodegeneration occurs, significantly altering the disease trajectory for affected individuals.

Our study aims to identify carriers of pathogenic and likely pathogenic *SOD1* variants in a large population dataset, in an era where a molecular therapy for *SOD1*-ALS is available and is currently being investigated in a clinical trial involving presymptomatic individuals. In addition, we aim to estimate the genetic prevalence of *SOD1*-ALS in the UK population, potentially expanding our understanding of *SOD1*-ALS epidemiology and informing current and future preventive and therapeutic strategies.

## Methods

### Study Population and Variant Detection

The UK Biobank (UKB) cohort is a large repository including longitudinal demographic, phenotypic and genomic data from more than 500,000 individuals between the ages of 40 and 69 years, recruited across the UK between 2006 and 2010^15^. Leveraging whole-exome sequencing (WES) data from 470,000 participants, we extracted *SOD1* coding variants with a minor allele frequency lower than 0.001. Variant annotation was performed using Annovar tool^16^, and synonymous, splicing and non-coding variants (in the 3’ and 5’ UTR) were excluded. Variants were classified as pathogenic, likely pathogenic, or of uncertain significance following American College of Medical Genetics and Genomics (ACMG) guidelines^17^ and according to reference databases specific to ALS, including ALSOD, SODCOD, and Project Mine^6,18,19^. We established a consensus classification by integrating evidence from multiple sources, with priority given to functionally validated variants with established genotype-phenotype correlations.

### Prevalence Estimation Methodology

To estimate the number of people affected by *SOD1*-ALS in the UK population, we employed a previously described modelling approach^20^, which incorporated:

1. *SOD1* carrier frequency in the UKB cohort, calculated by dividing the number of exomes with a pathogenic or likely pathogenic *SOD1* variant by the total number of sequences (n=470,000). Confidence intervals at 95% were calculated as previously described^20^.
2. Age-stratified population statistics from the UK Office of National Statistics^21^.
3. Distribution of age at onset of *SOD1*-ALS, derived from a cohort of N=1122 patients^12^. Since the p.Ala5Val variant was not identified in the UKB cohort, we excluded patients harbouring this variant from the age-at-onset distribution analysis to ensure our model accurately reflected the variant spectrum observed in our dataset.
4. Disease penetrance of *SOD1*-ALS, which was recently estimated to be 54% at a population level^5^.
5. Median disease survival, to adjust for mortality effects. Total *SOD1*-ALS median survival is reported to be 2.7 years^12^; however, since forms with the p.Ala5Val variants have a more aggressive disease course (survival 1.4 years), we considered the disease survival for non-p.Ala5Val *SOD1*-ALS as more representative (median survival 6.8 years)^12^.

Heterozygous p.Asp91Ala has been reported as a low-penetrance variant, and its pathogenic effect in *SOD1*-ALS is still debated. This variant appears to cause disease primarily in homozygous state or in compound heterozygosity with other pathogenic variants, with limited evidence for pathogenicity in heterozygous carriers^11^. For this reason, we performed two different estimates of *SOD1*-ALS genetic prevalence, with and without the p.Asp91Ala variant, to provide a comprehensive range of potential disease burden.

The new genetic estimate was compared to the clinical figure derived from literature data. Considering that *SOD1* mutations account for up to 20% of familial ALS (fALS), which represents 10% of all ALS cases, and 1–2% of sporadic ALS (sALS), which constitutes 90% of ALS cases, and given an ALS prevalence of 8 per 100,000 (range: 5–12 per 100,000)^22^, the prevalence of *SOD1*-ALS based on clinical observation would be 0.27 per 100,000.

### Phenotypic analysis

Age at recruitment, gender and ethnicity were collected from UKB baseline assessment data. To investigate potential disease manifestations among *SOD1* variant carriers, we implemented a comprehensive phenotyping approach using multiple data sources. We extracted medical diagnosis codes (ICD-10) from linked healthcare records and self-reported health conditions from participant questionnaires, along with the reported cause of death for deceased patients. We developed a hierarchical classification system to categorise symptom severity, where subjects with at least one code referred to the condition and/or to motor neuron degeneration signs and symptoms (**Supplementary Table 1**) were considered symptomatic. Symptoms were further stratified into strong, moderate, and weak evidence categories based on the specificity of ICD-10 codes to motor neuron disease (MND) and known *SOD1*-ALS manifestations.

### Statistical Analysis

All data were analysed using R Studio Version 4.4.3 (https://cran.rstudio.com/), and the statistical significance threshold was set to *p ≤ 0.05*. Continuous variables were compared between symptomatic and asymptomatic carriers using the Mann-Whitney U test. Categorical variables were compared between the same groups using Fisher’s exact test. Plots were generated using the ggplot2 package in R. Graphs representing proportions of symptomatic and asymptomatic individuals were generated using GraphPad Prism Version 10.

## Results

We identified a total of nineteen coding variants previously reported in the scientific literature and currently classified as pathogenic (N=9) and likely pathogenic (N=10) across 122 participants in the UKB cohort (**Fig. 1**, **Table 1**). Additionally, 535 individuals were found to carry the low-penetrance heterozygous p.Asp91Ala variant. Notably, no coding homozygous *SOD1* variants were identified in the cohort. None of these individuals harboured other *SOD1* variants.

**Figure 1.**
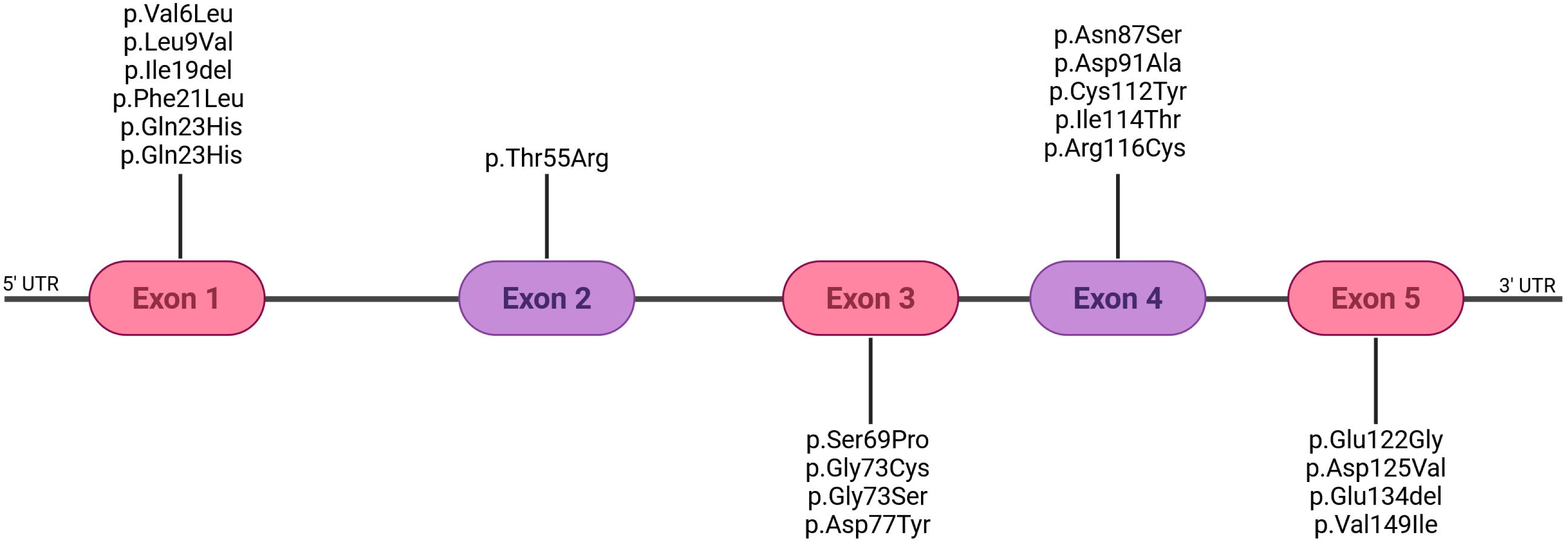
Schematic representation of the SOD1 gene showing the location of 20 distinct coding variants identified across 657 patients. Exons are depicted as colored boxes, and variants are marked along the gene structure according to their position.

**Table 1.**
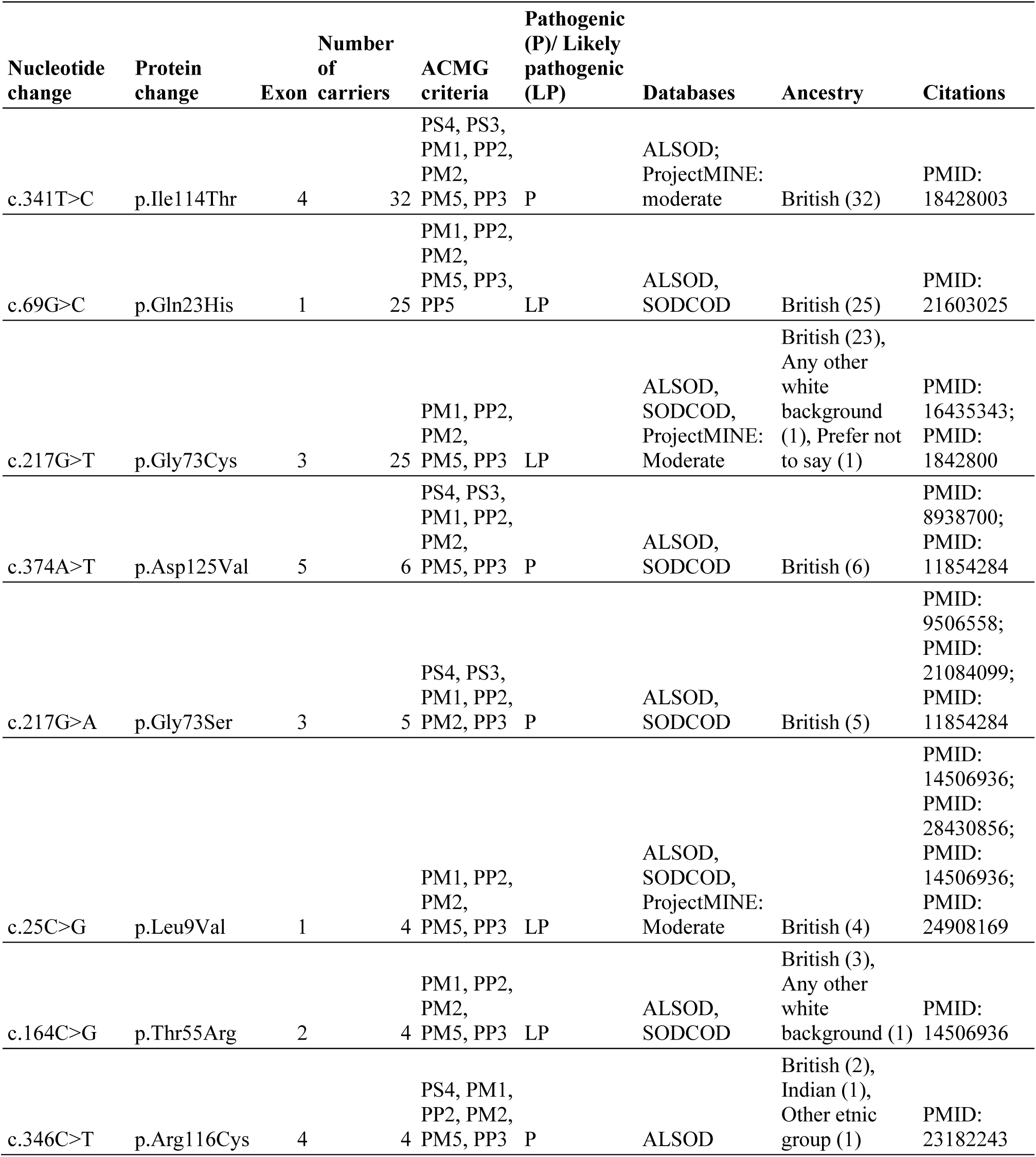
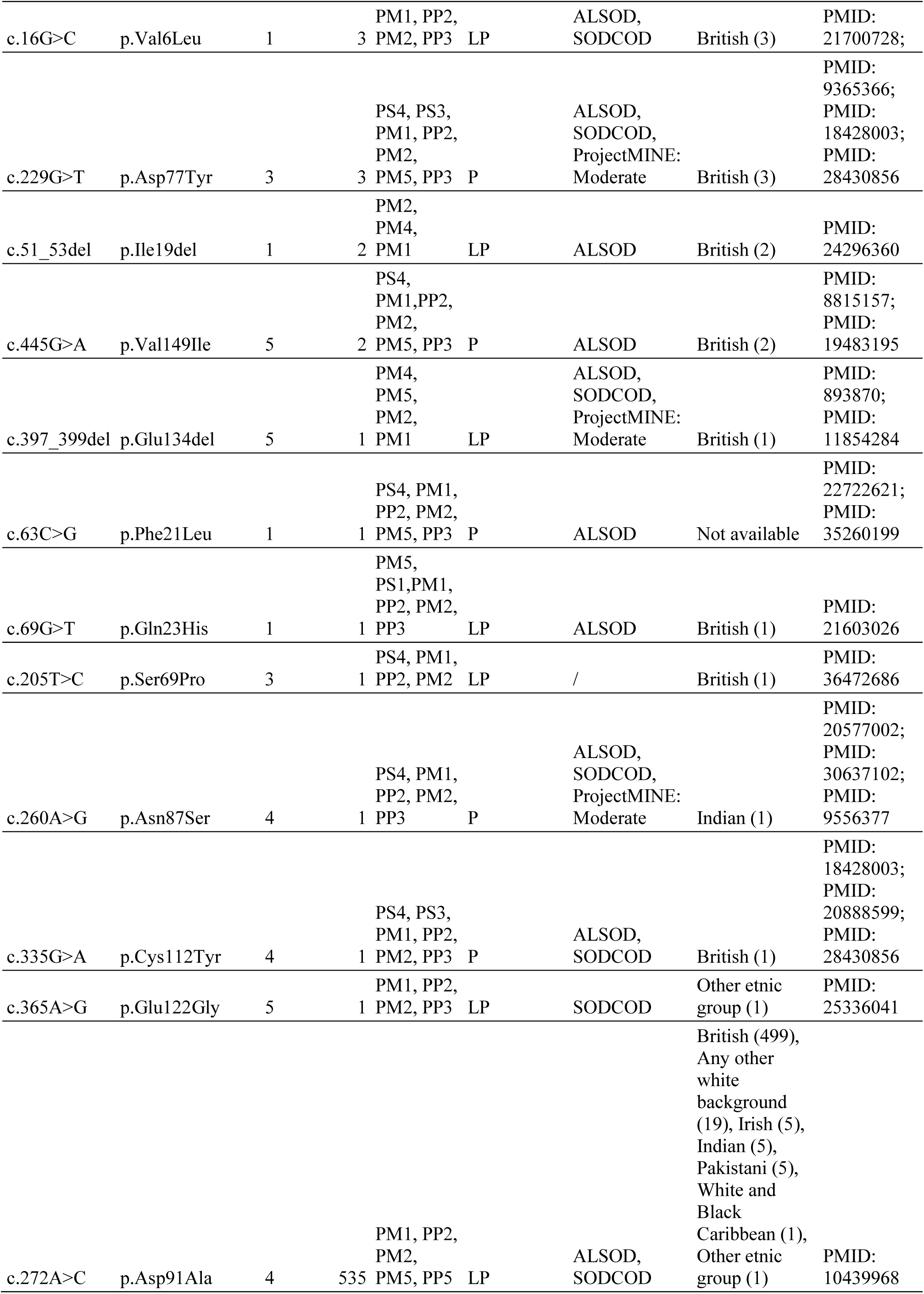
*SOD1* pathogenic and likely pathogenic variants identified in the UK Biobank cohort.

| Nucleotide change | Protein change | Exon | Number of carriers | ACMG criteria | Pathogenic (P)/ Likely pathogenic (LP) | Databases | Ancestry | Citations |
| --- | --- | --- | --- | --- | --- | --- | --- | --- |
| c.341T>C | p.Ile114Thr | 4 | 32 | PS4, PS3, PM1, PP2, PM2, PM5, PP3 | P | ALSOD; ProjectMINE: moderate | British (32) | PMID: 18428003 |
| c.69G>C | p.Gln23His | 1 | 25 | PM1, PP2, PM2, PM5, PP3, PP5 | LP | ALSOD, SODCOD | British (25) | PMID: 21603025 |
| c.217G>T | p.Gly73Cys | 3 | 25 | PM1, PP2, PM2, PM5, PP3 | LP | ALSOD, SODCOD, ProjectMINE: Moderate | British (23), Any other white background (1), Prefer not to say (1) | PMID: 16435343; PMID: 1842800 |
| c.374A>T | p.Asp125Val | 5 | 6 | PS4, PS3, PM1, PP2, PM2, PM5, PP3 | P | ALSOD, SODCOD | British (6) | PMID: 8938700; PMID: 11854284 |
| c.217G>A | p.Gly73Ser | 3 | 5 | PS4, PS3, PM1, PP2, PM2, PP3 | P | ALSOD, SODCOD | British (5) | PMID: 9506558; PMID: 21084099; PMID: 11854284 |
| c.25C>G | p.Leu9Val | 1 | 4 | PM1, PP2, PM2, PM5, PP3 | LP | ALSOD, SODCOD, ProjectMINE: Moderate | British (4) | PMID: 14506936; PMID: 28430856; PMID: 14506936; PMID: 24908169 |
| c.164C>G | p.Thr55Arg | 2 | 4 | PM1, PP2, PM2, PM5, PP3 | LP | ALSOD, SODCOD | British (3), Any other white background (1) | PMID: 14506936 |
| c.346C>T | p.Arg116Cys | 4 | 4 | PS4, PM1, PP2, PM2, PM5, PP3 | P | ALSOD | British (2), Indian (1), Other ethnic group (1) | PMID: 23182243 |
| c.16G>C | p.Val6Leu | 1 | 3 | PM1, PP2,<br>PM2, PP3 | LP | ALSOD,<br>SODCOD | British (3) | PMID:<br>21700728; |
| c.229G>T | p.Asp77Tyr | 3 | 3 | PS4, PS3,<br>PM1, PP2,<br>PM2,<br>PM5, PP3 | P | ALSOD,<br>SODCOD,<br>ProjectMINE:<br>Moderate | British (3) | PMID:<br>9365366;<br>PMID:<br>18428003;<br>PMID:<br>28430856 |
| c.51_53del | p.Ile19del | 1 | 2 | PM2,<br>PM4,<br>PM1 | LP | ALSOD | British (2) | PMID:<br>24296360 |
| c.445G>A | p.Val149Ile | 5 | 2 | PS4,<br>PM1,PP2,<br>PM2,<br>PM5, PP3 | P | ALSOD | British (2) | PMID:<br>8815157;<br>PMID:<br>19483195 |
| c.397_399del | p.Glu134del | 5 | 1 | PM4,<br>PM5,<br>PM2,<br>PM1 | LP | ALSOD,<br>SODCOD,<br>ProjectMINE:<br>Moderate | British (1) | PMID:<br>893870;<br>PMID:<br>11854284 |
| c.63C>G | p.Phe21Leu | 1 | 1 | PS4, PM1,<br>PP2, PM2,<br>PM5, PP3 | P | ALSOD | Not available | PMID:<br>22722621;<br>PMID:<br>35260199 |
| c.69G>T | p.Gln23His | 1 | 1 | PM5,<br>PS1,PM1,<br>PP2, PM2,<br>PP3 | LP | ALSOD | British (1) | PMID:<br>21603026 |
| c.205T>C | p.Ser69Pro | 3 | 1 | PS4, PM1,<br>PP2, PM2 | LP | / | British (1) | PMID:<br>36472686 |
| c.260A>G | p.Asn87Ser | 4 | 1 | PS4, PM1,<br>PP2, PM2,<br>PP3 | P | ALSOD,<br>SODCOD,<br>ProjectMINE:<br>Moderate | Indian (1) | PMID:<br>20577002;<br>PMID:<br>30637102;<br>PMID:<br>9556377 |
| c.335G>A | p.Cys112Tyr | 4 | 1 | PS4, PS3,<br>PM1, PP2,<br>PM2, PP3 | P | ALSOD,<br>SODCOD | British (1) | PMID:<br>18428003;<br>PMID:<br>20888599;<br>PMID:<br>28430856 |
| c.365A>G | p.Glu122Gly | 5 | 1 | PM1, PP2,<br>PM2, PP3 | LP | SODCOD | Other ethnic<br>group (1) | PMID:<br>25336041 |
| c.272A>C | p.Asp91Ala | 4 | 535 | PM1, PP2,<br>PM2,<br>PM5, PP5 | LP | ALSOD,<br>SODCOD | British (499),<br>Any other<br>white<br>background<br>(19), Irish (5),<br>Indian (5),<br>Pakistani (5),<br>White and<br>Black<br>Caribbean (1),<br>Other ethnic<br>group (1) | PMID:<br>10439968 |

Finally, 66 additional previously unreported variants were identified in 401 individuals. Among them, 36 were pathogenic or likely pathogenic, while the remaining were classified as variants of unknown significance. Considering the absence of clinical or functional evidence of pathogenicity, we excluded them from further analyses (**Supplementary Table 2**).

The carrier frequency for pathogenic and likely pathogenic *SOD1* variants in UKB was calculated as 1 out of 3,852 individuals (95% CI: 1 in 3,241 to 1 in 4,627). When including the heterozygous p.Asp91Ala variant, the carrier frequency increased substantially to 1 in 715 individuals (95% CI: 1 in 677 to 1 in 755). The carrier frequency of the heterozygous p.AspD91Ala variant in the UKBB is 1 in 878 (95% CI: 1 in 810 to 1 in 960).

To estimate the prevalence of *SOD1*-ALS in the UK population, we applied a disease modelling approach using age-at-onset data from 598 patients with *SOD1*-ALS, excluding the p.Ala5Val variant^12^. The number of people with *SOD1*-ALS was estimated at 1.1 cases per 100,000 individuals, which is approximately four times higher than the prevalence derived from clinical observation (0.27 per 100,000) (**Fig.2A**). When including the low-penetrance heterozygous p.Asp91Ala variant and using age-at-onset data from 667 patients^12^, the prevalence of *SOD1*-ALS increased drastically to 5.7 cases per 100,000 individuals (**Fig. 2B**).

**Figure 2.**
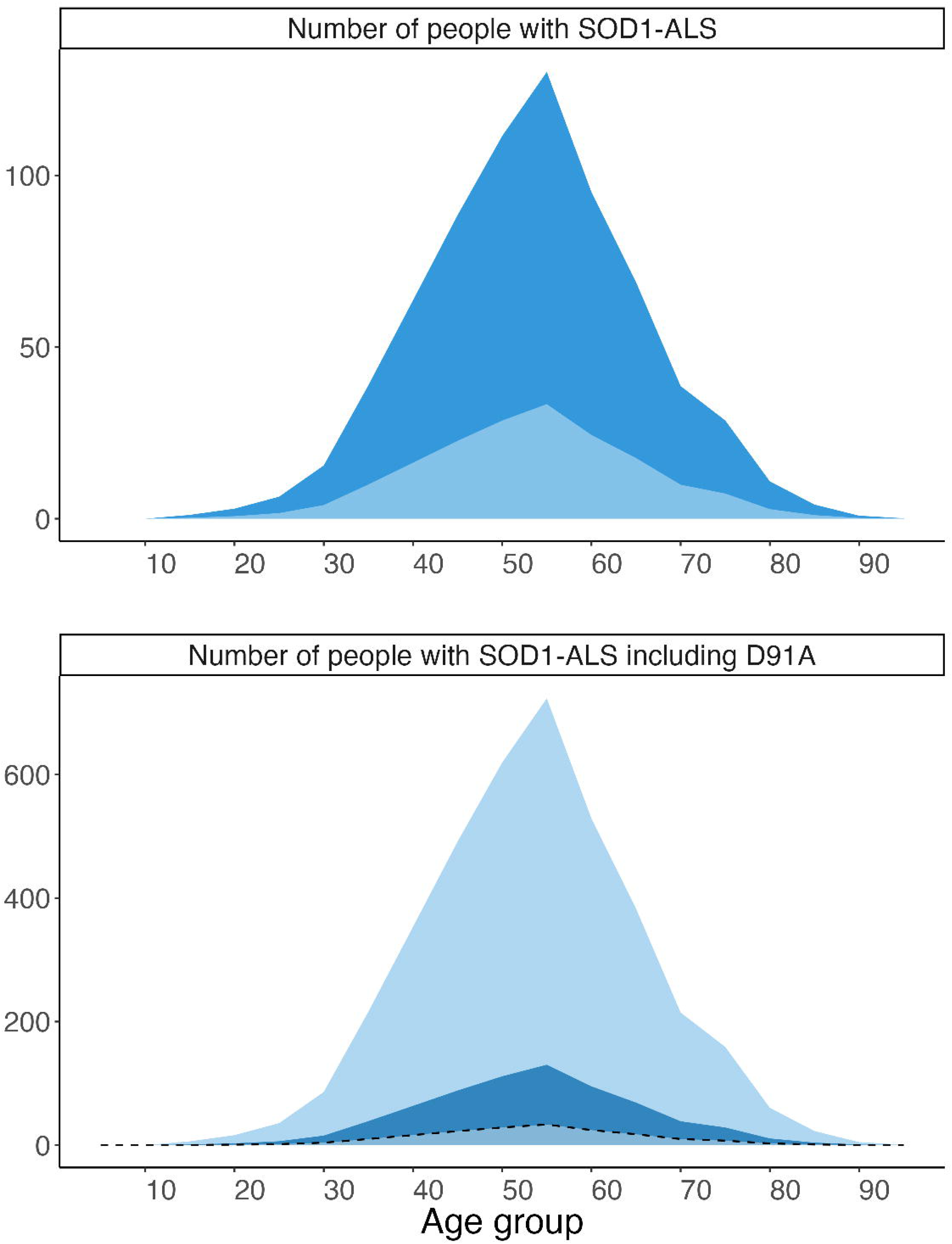
Estimated number of people with *SOD1*-ALS due to pathogenic and likely pathogenic variants in the UK Biobank cohort. **A.** Prevalence estimates (dark blue area) are calculated using a carrier frequency of 1/3852 and do not include carriers of the p.Asp91Ala variant, and are compared to clinical prevalence from the literature (light blue area). **B.** Prevalence estimates (light blue area) are calculated using a carrier frequency of 1/715, including also carriers of the p.Asp91Ala variant, and are compared to the newly estimated prevalence (A, dark blue area) and to the clinical prevalence from the literature (light blue area **surrounded by a dashed line**). The UK population count by age is multiplied by the disease allele frequency of SOD1 variants and the age of onset distribution of SOD1-ALS, and corrected for median survival. Penetrance is assumed to be 50%. X-axis: The age bins are 5 years each; y-axis: estimated number of affected individuals.

Demographic analysis of the 657 identified *SOD1* carriers of pathogenic and likely pathogenic variants revealed a mean age at recruitment of 58 [50.5-63] years, with 45.8% being female (**Fig.3C**). The majority (97.3%) were of European ancestry.

**Figure 3.**
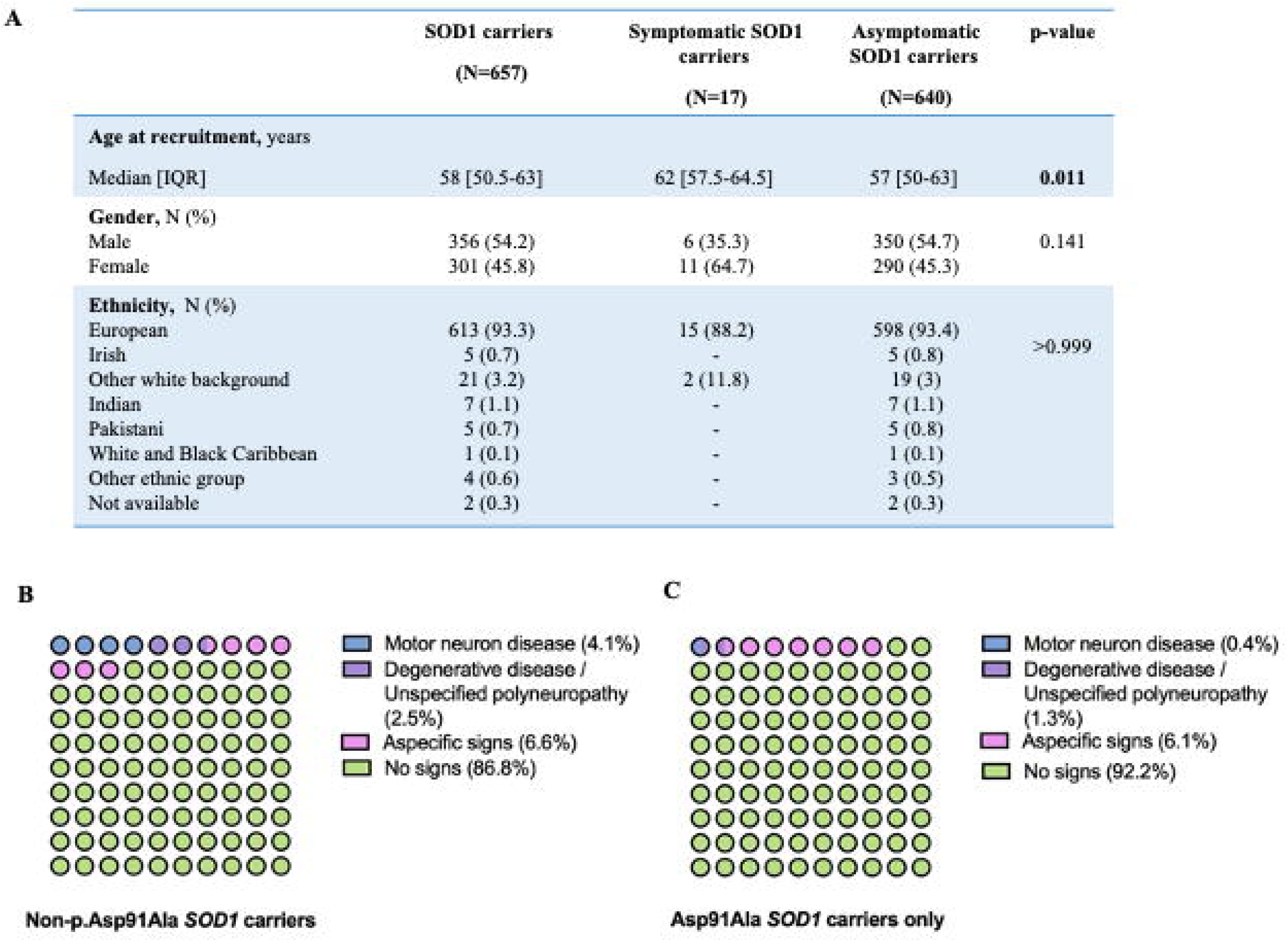
*SOD1* carriers according to symptomatic state. **A.** Age at recruitment, gender and ethnicity of carriers of *SOD1* pathogenic and likely pathogenic variants in the UK Biobank cohort, according to the presence of ICD-10 codes compatible with ALS. Only carriers with strong or moderate evidence of symptoms were considered symptomatic **B-C.** Waffle plots representing the proportion of symptomatic patients among *SOD1* non-p.Asp91Ala carriers (B) and in p.Asp91Ala carriers only (C).

Clinical phenotyping identified 17 symptomatic individuals (2.6%) in the overall cohort. Definite MND diagnosis (strong evidence) was observed in N=7 (1.1%) individuals (4.1% of the non-p.Asp91Ala carriers and only 0.4% among the p.Asp91Ala carriers), while non-specific neurodegenerative conditions or polyneuropathy (moderate evidence) were detected in N=10 (1.5%) individuals (N=3, 2.5% of non-Asp91Ala and N=7, 1.3% of p.Asp91Ala carriers), and non-specific neurological symptoms (weak evidence) were detected in N=40, 6.1% of subjects (**Fig.3B-C**). For subsequent analyses, only carriers with strong or moderate evidence of symptoms were considered symptomatic.

Comparing symptomatic versus asymptomatic carriers, a significant difference was found in age at recruitment (62 [57.5-64.5] vs. 57 [50-63] years, **p=0.011**), which was still present when considering only the non-p.Asp91Ala carriers, but no significant differences in sex or ancestry distribution were observed (**Fig.3A**).

## Discussion

Our comprehensive analysis of *SOD1* variants in the UKB cohort reveals a substantial discrepancy between genetic and clinical prevalence of *SOD1*-ALS, with important implications for screening strategies and therapeutic interventions. The identification of 122 carriers of pathogenic or likely pathogenic *SOD1* variants among 470,000 participants indicates a carrier frequency of 1 in 3,852, which is considerably higher than anticipated based on clinical diagnoses alone. This finding is consistent with previous work on the genetic prevalence of short tandem repeat expansions causative for neurological diseases in large genomic cohorts^20,23^. In particular, *C9orf72*-related ALS was estimated to be over two times higher than the prevalence from clinical observation.

This discrepancy can be attributed to several factors. First, incomplete penetrance appears to play a significant role, with only 2.6% of carriers displaying neurological symptoms at enrollment. Second, phenotypic heterogeneity likely contributes to underdiagnosis, as some *SOD1*-ALS cases present with atypical features or slow progression that may not prompt neurological evaluation or accurate diagnosis^24^. The longitudinal design of UKB, along with the addition of increasingly detailed clinical information, would enable a better understanding of age-dependent penetrance and disease expressivity in *SOD1*-ALS.

The four-fold difference between genetic prevalence (1.1 per 100,000) and clinical prevalence (0.27 per 100,000) has substantial clinical implications. This gap suggests that many individuals who could benefit from SOD1-directed therapies remain undiagnosed. The recent approval of tofersen for *SOD1*-ALS patients provides unprecedented opportunities for intervention, potentially modifying the disease course if administered early. Furthermore, emerging evidence from longitudinal studies of pre-symptomatic carriers indicates that biomarkers like plasma NfL can identify individuals approaching phenoconversion, creating a window for preventive treatment before irreversible neurodegeneration occurs^13^.

Our finding that the heterozygous p.Asp91Ala variant may cause phenotypes compatible with MND in a limited portion of carriers supports the contribution of this variant also in heterozygous state, as previously observed^3^. However, the low rate of symptomatic individuals among p.Asp91Ala carriers acknowledges the role of p.Asp91Ala as a low-penetrance allele requiring additional genetic or environmental factors to manifest disease, consistent with conclusions from previous family studies^25^. Nevertheless, the reduction in serum NfL levels following tofersen administration^26^ supports the eligibility of symptomatic p.Asp91Ala carriers for this molecular therapy.

Similarly, the p.Ser69Pro has been demonstrated so far as causative for early-onset ALS only in homozygosis^9^. In this study, the same variant was identified in heterozygosis in a single subject recruited at the age of 56, who died of MND. Thus, this finding confirms the association between monoallelic defects, initially described as recessive alleles, and late-onset clinical manifestations^10^.

Our study is not exempt from limitations. First of all, the UKB cohort represents individuals aged 40-69 at recruitment, potentially missing younger and older onset cases. Additionally, the ICD-10 codes might not capture the full clinical details of individuals at any given time, being recorded and updated only if patients are admitted to the hospital, irrespective of the presence of symptoms. Moreover, the follow-up period may be insufficient to capture late-onset disease manifestations. No information is available about the family history of neuromuscular disorders, nor have instrumental investigations been performed to identify pre- or subclinical motor neuron damage or abnormal muscle activity. Secondly, our analysis mainly focused on previously reported *SOD1*-causative single nucleotide variants (SNVs) in coding regions, excluding molecular defects lacking a conclusive demonstration of pathogenicity as well as structural and non-coding variants. In this study, we have reported a list of previously unreported SNVs, which we did not consider in the genetic prevalence calculation because of a lack of evidence of pathogenicity. We acknowledge that including those variants in the disease modelling would further increase the prevalence estimates. At the same time, a recent study reporting an elevated frequency of *SOD1* variants in gnomAD dataset points towards a possible reduced penetrance of *SOD1*-ALS in the general population^27^, mitigating the effective clinical impact of *SOD1* variants identified in large genomic datasets. In addition, age at disease onset and penetrance are highly variable across different *SOD1* variants, while we considered an overall penetrance of 54%, as previously reported ^5^. Finally, even if other ancestries are reported, the UKB cohort is predominantly composed of participants of European ancestry, which might under-represent the genetic complexity of the worldwide population.

These findings support the implementation of more systematic genetic screening approaches for *SOD1* variants, particularly in the context of a family history of ALS or unexplained neurological symptoms. The availability of effective targeted therapy transforms the risk-benefit calculation for genetic testing, as identification of presymptomatic carriers now offers tangible clinical benefits through potential early intervention.

Future research should focus on identifying additional genetic and environmental modifiers affecting *SOD1* variant penetrance and developing improved predictive models of phenoconversion risk. Implementation studies examining the clinical utility and cost-effectiveness of broader genetic screening programs would also provide valuable guidance for clinical practice and to support policy decisions in national health services.

In conclusion, our study reveals that *SOD1*-related ALS is significantly more prevalent at the genetic level than previously recognised through clinical diagnosis alone. Given that *SOD1*-ALS is currently the only genetic form of ALS with an approved molecular therapy^28^, and that early intervention may be critical for treatment efficacy, these results underscore the importance of identifying at-risk individuals before symptom onset or in early disease stages.

### Ethics approval

All research activities followed protocols approved by the Research Ethics Committee (UKBB reference: 16/NW/0274), with all study participants providing informed consent.

## Data availability statement

All data produced in the present study are available upon reasonable request to the corresponding authors.

## Acknowledgments

This work was promoted within the European Reference Network (ERN) for Rare Neuromuscular Diseases. We thank the Associazione Centro Dino Ferrari for its support. The PNC “Hub Life Science-Diagnostica Avanzata (HLS-DA), PNC-E3-2022-23683266– CUP: C43C22001630001” is funded by the Italian Minister of Health. The support of the Italian Ministry of Education and Research (MUR) “Dipartimenti di Eccellenza Program 2023–2027” - Dept of Pathophysiology and Transplantation, University of Milan to DR, SC and GPC is gratefully acknowledged.

## Supplementary material

**Supplementary Table 1. ICD-10 codes compatible with motor neuron disease with strong, moderate and weak evidence.**

**Supplementary Table 2.** *SOD1* coding variants not reported in the literature and with no conclusive demonstration of pathogenicity.

