## Supplementary Table 1 for "High Prevalence of *SOD1* Pathogenic Variants in the UK Biobank: Implications for Early Intervention in ALS"

**Supplementary table 1. ICD-10 codes compatible with motor neuron disease with strong, moderate and weak evidence.**

**STRONG**

G12.1 Other inherited spinal muscular atrophy

G12.2 Motor neuron disease

G12.8 Other spinal muscular atrophies and related syndromes

**MODERATE**

G31.8 Other specified degenerative diseases of nervous system

G31.9 Degenerative disease of nervous system, unspecified

G32.8 Other specified degenerative disorders of nervous system in diseases classified elsewhere

G62.9 Polyneuropathy, unspecified

G62.8 Other specified polyneuropathies

**WEAK**

G31.0 Circumscribed brain atrophy

G31.1 Senile degeneration of brain, not elsewhere classified

G60.3 Idiopathic progressive neuropathy

G60.8 Other hereditary and idiopathic neuropathies

G60.9 Hereditary and idiopathic neuropathy, unspecified

G63.6 Polyneuropathy in other musculoskeletal disorders

G63.8 Polyneuropathy in other diseases classified elsewhere

G64 Other disorders of peripheral nervous system

R25.2 Cramp and spasm

R25.3 Fasciculation

R25.8 Other and unspecified abnormal involuntary movements

R26.1 Paralytic gait

R26.2 Difficulty in walking, not elsewhere classified

R26.3 Immobility

R26.8 Other and unspecified abnormalities of gait and mobility

R29.2 Abnormal reflex

R29.6 Tendency to fall, not elsewhere classified

R29.8 Other and unspecified symptoms and signs involving the nervous and musculoskeletal systems

R47.0 Dysphasia and aphasia

R47.1 Dysarthria and anarthria

R47.8 Other and unspecified speech disturbances

R49.0 Dysphonia

R49.1 Aphonia

R49.8 Other and unspecified voice disturbances
