## Supplementary table 2 for "High Prevalence of *SOD1* Pathogenic Variants in the UK Biobank: Implications for Early Intervention in ALS"

**Supplementary table 2. *SOD1* coding variants in UK Biobank not reported in the literature and with no conclusive demonstration of pathogenicity.**

| **Nucleotide Change** | **Protein Change** | **Exon** | **UKBB frequency** | **ACMG Criteria** | **P/LP/VUS** | **Variant Type** | **Databases** |
| --- | --- | --- | --- | --- | --- | --- | --- |
| c.8C>G | p.Thr3Arg | 1 | 2 | PM1, PP2, PM2 | VUS | Nonsynonymous SNV |  |
| c.22G>A | p.Val8Met | 1 | 1 | PM1, PP2, PM2, PM5, PP3 | LP | Nonsynonymous SNV |  |
| c.34G>A | p.Asp12Asn | 1 | 1 | PM1, PP2, PM2, PM5 | LP | Nonsynonymous SNV |  |
| c.36C>A | p.Asp12Glu | 1 | 2 | PM1, PP2, PM2, PM5 | LP | Nonsynonymous SNV |  |
| c.40C>T | p.Pro14Ser | 1 | 1 | PM1, PP2, PM2 | VUS | Nonsynonymous SNV |  |
| c.47A>G | p.Gln16Arg | 1 | 25 | PM1, PP2, PM2 | VUS | Nonsynonymous SNV |  |
| c.48G>C | p.Gln16His | 1 | 4 | PM1, PP2, PM2 | VUS | Nonsynonymous SNV |  |
| c.50G>T | p.Gly17Val | 1 | 1 | PM1, PP2, PM2, PM5, PP3 | LP | Nonsynonymous SNV |  |
| c.70A>C | p.Lys24Gln | 1 | 1 | PM1, PP2, PM2 | VUS | Nonsynonymous SNV |  |
| c.74A>G | p.Glu25Gly | 2 | 2 | PM2, PP2 | VUS | Nonsynonymous SNV |  |
| c.77G>A | p.Ser26Asn | 2 | 1 | PM2, PP2 | VUS | Nonsynonymous SNV |  |
| c.82G>A | p.Gly28Arg | 2 | 3 | PM2, PM1, PP2 | VUS | Nonsynonymous SNV |  |
| c.86C>G | p.Pro29Arg | 2 | 9 | PM2, PM1, PP2 | VUS | Nonsynonymous SNV |  |
| c.92A>G | p.Lys31Arg | 2 | 2 | PM2, PM1, PP2 | VUS | Nonsynonymous SNV |  |
| c.92A>T | p.Lys31Met | 2 | 1 | PM2, PM1, PP2 | VUS | Nonsynonymous SNV |  |
| c.136T>G | p.Phe46Val | 2 | 1 | PM1, PP2, PM2, PP3, PM5 | LP | Nonsynonymous SNV |  |
| c.138C>G | p.Phe46Leu | 2 | 2 | PM1, PP2, PM2, PM5, PP3 | LP | Nonsynonymous SNV |  |
| c.164C>T | p.Thr55Ile | 2 | 2 | PM1, PP2, PM2, PP3, PM5 | LP | Nonsynonymous SNV |  |
| c.173G>C | p.Cys58Ser | 3 | 1 | PM2, PM1, PP2, PP3 | VUS | Nonsynonymous SNV |  |
| c.181G>A | p.Ala61Thr | 3 | 1 | PM2, PM1, PP2, PP3 | VUS | Nonsynonymous SNV |  |
| c.182C>G | p.Ala61Gly | 3 | 1 | PM2, PM1, PP2, PP3 | VUS | Nonsynonymous SNV |  |
| c.182C>T | p.Ala61Val | 3 | 2 | PM2, PM1, PP2, PP3 | VUS | Nonsynonymous SNV |  |
| c.216C>A | p.His72Gln | 3 | 2 | PM1, PP2, PM2, PM5, PP3 | LP | Nonsynonymous SNV |  |
| c.221G>A | p.Gly74Glu | 3 | 1 | PM1, PP2, PM2, PP3 | LP | Nonsynonymous SNV |  |
| c.229G>A | p.Asp77Asn | 3 | 1 | PM1, PP2, PM2, PM5, PP3 | LP | Nonsynonymous SNV |  |
| c.274A>G | p.Lys92Glu | 4 | 1 | PM1, PP2, PM2 | VUS | Nonsynonymous SNV |  |
| c.276A>T | p.Lys92Asn | 4 | 1 | PM1, PP2, PM2 | VUS | Nonsynonymous SNV |  |
| c.277G>C | p.Asp93His | 4 | 2 | PM1, PP2, PM2 | VUS | Nonsynonymous SNV |  |
| c.278A>G | p.Asp93Gly | 4 | 7 | PM1, PP2, PM2 | VUS | Nonsynonymous SNV |  |
| c.284T>C | p.Val95Ala | 4 | 9 | PM2, PM1, PP2, PP3 | LP | Nonsynonymous SNV | ALSOD, SODCOD |
| c.287C>T | p.Ala96Val | 4 | 5 | PM1, PP2, PM2, PM5, PP3 | LP | Nonsynonymous SNV | SODCOD |
| c.290A>T | p.Asp97Val | 4 | 1 | PM1, PP2, PM2, PM5 | LP | Nonsynonymous SNV |  |
| c.298A>G | p.Ile100Val | 4 | 8 | PM1, PP2, PM2 | VUS | Nonsynonymous SNV | LOVD (VUS), ALSOD (benign/neutral), SODCOD |
| c.299T>C | p.Ile100Thr | 4 | 2 | PM2, PM1, PP2, PP3 | LP | Nonsynonymous SNV |  |
| c.307T>A | p.Ser103Thr | 4 | 8 | PM1, PP2, PM2 | VUS | Nonsynonymous SNV |  |
| c.314T>C | p.Ile105Thr | 4 | 1 | PM1, PP2, PM2, PM5, PP3 | LP | Nonsynonymous SNV |  |
| c.331C>G | p.His111Asp | 4 | 3 | PM1, PP2, PM2 | VUS | Nonsynonymous SNV |  |
| c.335G>C | p.Cys112Ser | 4 | 1 | PM1, PP2, PM2, PM5 | LP | Nonsynonymous SNV |  |
| c.347G>A | p.Arg116His | 4 | 9 | PS4, PM1, PP2, PM2, PM5, PP3 | P | Nonsynonymous SNV |  |
| c.382G>A | p.Gly128Ser | 5 | 1 | PM2, PM1, PP2, PP3 | LP | Nonsynonymous SNV |  |
| c.385A>G | p.Lys129Glu | 5 | 2 | PM2, PM1, PP2, PP3 | LP | Nonsynonymous SNV |  |
| c.388G>A | p.Gly130Ser | 5 | 2 | PM2, PM1, PP2, PP3 | LP | Nonsynonymous SNV |  |
| c.391G>A | p.Gly131Arg | 5 | 1 | PM2, PM1, PP2, PP3 | LP | Nonsynonymous SNV |  |
| c.392G>A | p.Gly131Glu | 5 | 4 | PM2, PM1, PP2, PP3 | LP | Nonsynonymous SNV |  |
| c.407C>T | p.Thr136Ile | 5 | 38 | PM1, PP2, PM2 | VUS | Nonsynonymous SNV |  |
| c.421G>C | p.Ala141Pro | 5 | 1 | PM2, PM1, PP2, PP3 | LP | Nonsynonymous SNV |  |
| c.430C>T | p.Arg144Cys | 5 | 4 | PM2, PM1, PP2, PP3 | LP | Nonsynonymous SNV |  |
| c.431G>A | p.Arg144His | 5 | 2 | PM2, PM1, PP2, PP3 | LP | Nonsynonymous SNV |  |
| c.448A>G | p.Ile150Val | 5 | 1 | PM1, PP2, PM2, PM5, PP5 | LP | Nonsynonymous SNV | SODCOD |
| c.457G>A | p.Ala153Thr | 5 | 2 | PM1, PP2, PM2 | VUS | Nonsynonymous SNV | ProjectMINE (moderate) |
| c.58delA | p.Asn20Ilefs*11 | 1 | 3 | PVS1, PM2 | LP | Frameshift deletion |  |
| c.240_250del | p.His81Leufs*7 | 4 | 3 | PVS1, PM2 | LP | Frameshift deletion |  |
| c.120_121insA | p.Glu41Argfs*10 | 2 | 1 | PVS1, PM2 | LP | Frameshift insertion |  |
| c.421dupG | p.Ala141Glyfs*23 | 5 | 1 | PVS1, PM2 | LP | Frameshift insertion |  |
| c.2T>C | p.Met1? | 1 | 1 | PM2, PVS1 | VUS | Startloss |  |
| c.98G>A | p.Trp33* | 2 | 2 | PVS1, PM2 | LP | Stopgain |  |
| c.323C>G | p.Ser108* | 4 | 1 | PVS1, PM2 | LP | Stopgain |  |
| c.464A>C | p.*155Serext*6 | 5 | 8 | PM2,PM4 | VUS | Stoploss | projectMINE (high) |
| c.465A>T | p.*155Tyrext*6 | 5 | 2 | PM2,PM4 | VUS | Stoploss |  |
| c.72+1G>A |  | 1 | 1 | PVS1, PM2 | LP | Splicing |  |
| c.72+2C>T |  | 1 | 1 | PVS1, PM2 | LP | Splicing |  |
| c.73-2A>G |  | 2 | 2 | PVS1, PM2 | LP | Splicing |  |
| c.357+1insT |  | 4 | 16 | PM2,PP3 | VUS | Splicing |  |
| c.328G>T |  | 4 | 40 | PM1, PP2, PM2 | VUS | Nonsynonymous SNV | ALSOD, SODCOD |
| c.59A>G | p.Asn20Ser | 1 | 81 | PM1, PP2, PM2, BP6 | VUS | Nonsynonymous SNV | LOVD (LB), ALSOD, SODCOD, projectMINE (moderate) |
| c.289G>A | p.Asp97Asn | 4 | 4 | PM1, PP2, PM2, PM5 | LP | Nonsynonymous SNV | ALSOD |
